# No evidence that analgesic use after COVID-19 vaccination negatively impacts antibody responses

**DOI:** 10.1101/2022.10.14.22281103

**Authors:** Bonnie J. Lafleur, Lisa White, Michael D. Dake, Janko Z. Nikolich, Ryan Sprissler, Deepta Bhattacharya

## Abstract

Uptake of mRNA vaccines, especially booster immunizations, against COVID-19 has been lower than hoped, perhaps in part due to their reactogenicity. Analgesics might alleviate symptoms associated with vaccination, but studies to measure their impact on immune responses have been limited to relatively small cohorts. We semi-quantitatively measured antibody responses following COVID-19 vaccination in 2354 human participants surveyed about analgesic use. Participants who used non-steroidal anti-inflammatory drugs (NSAIDs) or acetaminophen after vaccination showed elevated antibody levels against the receptor binding domain of Spike protein relative to those who did not use analgesics. This pattern was observed for both mRNA-1273 and BNT162b2 and across age groups. Participants who used analgesics more frequently reported fatigue, muscle aches, and headaches than those who did not use painkillers. Amongst participants who reported these symptoms, we observed no statistically significant differences in antibody levels irrespective of analgesic use. These data suggest that antibody levels are elevated as a function of symptoms and inflammatory processes rather than painkiller use per se. Taken together, we find no evidence that analgesic use reduces antibody responses after COVID-19 vaccination. Recommendation of their use to alleviate symptoms might improve uptake of booster immunizations.

## Introduction

Messenger RNA (mRNA) vaccines against SARS-CoV-2, the virus that causes COVID-19, showed high efficacy against symptomatic illness caused by the ancestral strain (1, 2). Yet in part due to viral evolution and escape from neutralizing antibodies and in part due to waning of protective antibody production, the initial effectiveness of these vaccines has decreased from their peaks, especially against mild symptomatic infections (3). In response, vaccines updated to more closely match the circulating lineages have been made available in hopes of restoring high levels of protection against SARS-CoV-2 infections and COVID-19. Studies of neutralizing antibody levels and real-world effectiveness showed the benefit of bivalent boosters updated to match the BA.4-5 lineages at protecting against variants of concern (4–12). Yet uptake was poor, as only ~30% of eligible adults in the United States received the bivalent booster (13). Uptake is also likely to be low for the latest monovalent booster that has been updated to match the XBB1.5 lineage. New public campaigns and strategies will be needed to improve the uptake over what was observed previously.

The reactogenicity of mRNA vaccines might negatively impact willingness to receive booster immunizations (14). While serious side effects following vaccination are rare, mRNA vaccines frequently lead to mild local and systemic adverse events such as injection site pain, lymphadenopathy, myalgia, and fever (15, 16). Over-the-counter analgesics might mitigate some of these mild adverse events, but animal models of SARS-CoV-2 infection have shown that non-steroidal anti-inflammatory drugs (NSAIDs) substantially reduce antiviral antibody responses (17). It remains unclear whether these same inhibitory effects are seen in humans after mRNA vaccination and how different classes of painkillers impact the antibody response. Several prior studies have suggested minimal or even positive impacts of antipyretic use on antibody responses (18–20). Further analysis of their impact, independent of symptoms and inflammation, as well as subgroup analyses on variables such as age, vaccine type, and the class of analgesics, would be valuable to help interpret these findings.

## Materials and methods

### Human subjects

All human subject work was approved by the University of Arizona IRB and was conducted in accordance with all federal, state, and local regulations and guidelines under the protocols 1510182734 and 1410545697A048. Subjects under 18 years of age were excluded. Subjects were recruited via public announcement and website registration as part of the University of Arizona Antibody Testing Initiative. Following website registration, subjects were ascertained to be afebrile and without COVID-19 symptoms based on questionnaire, were consented, and bled. Blood was centrifuged at multiple sites across Arizona. For all subjects, venous blood was obtained by venipuncture into SST Vacutainer tubes (Becton Dickinson, Sunnyvale, CA, cat. #367988), serum separated by centrifugation at 1,200 rpm and sent to the central processing laboratory within 4 h.

### Statistical Methods

Two-sided t-test statistics within one-way analysis of variance were used to compare pairwise RBD ELISA OD450 means between pain reliever groups at a significance level of 0.05. Logistic regression, using likelihood ratio chi-square test statistics, with significance level of 0.05, were used to compare the proportion of participants experiencing symptoms by pain reliever groups. Comparisons of symptoms by seropositivity and seronegativity is descriptive, due to the small sample size of the seronegative participants. All analysis were performed using the R programming language (version 4.3.1).

### ELISAs

Mammalian RBD was purchased from GenScript (catalog # Z03483). S2 of the was purchased from Sino Biological (catalog # 40590-V08B). Enzyme-linked immunosorbent assay (ELISA) was performed as described (21). Plates were blocked with 1% non-fat dehydrated milk extract (Santa Cruz Biotechnology #sc-2325) in sterile PBS (Fisher Scientific Hyclone PBS #SH2035,) for 1 h, washed with PBS containing 0.05% Tween-20, and overlaid with a 1:40 dilution of serum for 60 min. Plates were then washed and incubated for 1hr in 1% PBS and milk containing an anti-human Pan-Ig HRP conjugated antibody (Jackson ImmunoResearch catalog 109-035-064) at a concentration of 1:2000 for 1 h. Plates were washed with PBS-Tween solution followed by PBS wash. To develop, plates were incubated in tetramethylbenzidine prior to quenching with 2N H2SO4. Plates were read for 450nm absorbance on CLARIOstar Plus from BMG Labtech. Samples with OD630 values greater than 0.05 were re-run. Every plate contained at least 32 seronegative controls and either CR3022 or HM3128 (Creative Diagnostics) monoclonal antibodies as a positive control for RBD or S2, respectively. This assay has received Emergency Use Authorization from the US Food and Drug Administration (ID 201116).

### Thresholds for SARS-CoV-2 Spike protein RBD seropositivity

Diagnostic seropositivity based on RBD and S2 ELISA OD450 levels, is defined using an RBD positivity threshold of greater than or equal to 0.4160. If the OD450 of RBD antibodies is below 0.1480, samples are considered seronegative. When the OD450 of RBD antibodies is between 0.1480 and 0.4160, samples are evaluated for antibodies against the S2 region of Spike protein. Any such samples with an S2 OD450 antibody value greater than 0.195 are considered seropositive, while all others are diagnostically seronegative. The diagnostic workflow is shown below.

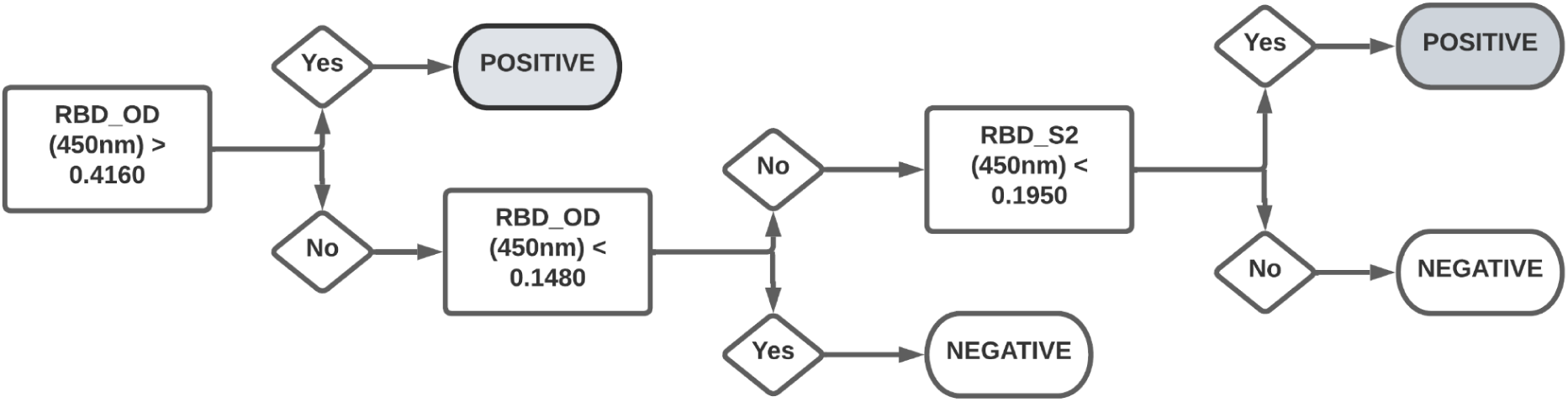

## Results

We examined survey results for analgesic use from March 15, 2021 through March 22, 2022 that included 2,354 vaccinated individuals that were part of a large statewide antibody testing initiative run by The University of Arizona (**Table 1**). Antibodies against the SARS-CoV-2 Spike receptor binding domain, self-reported vaccination information, and reported analgesic use within 48-hours after either two-dose mRNA COVID-19 vaccines (mRNA-1273 or BNT162b2) were analyzed. A one-way Analysis of Variance (ANOVA), using the t-test statistic, showed statistically significantly lower RBD-specific antibody levels in those who did not take an analgesic (n = 1,184) relative to those that took either NSAIDs or acetaminophen (**Fig. 1A**, p = 0.0001 for both). There was no statistically significant difference detected between the NSAID (n = 679) and acetaminophen (n = 491) analgesic groups (**Fig. 1A**, p = 0.9332). Thus analgesic use was associated with higher, rather than lower levels of anti-Spike antibodies. These differences were not modified by age group or vaccine, as determined by evaluation of the interaction effect between these variables and analgesic group (**Fig. 1B-C**, p = 0.0834 and p = 0.0819, respectively). For reasons that are unclear, and therefore results are not shown, those who did not answer the painkiller questionnaire (n = 1489) had statistically significantly higher RBD levels than those that did respond to the survey.

**Figure 1.**
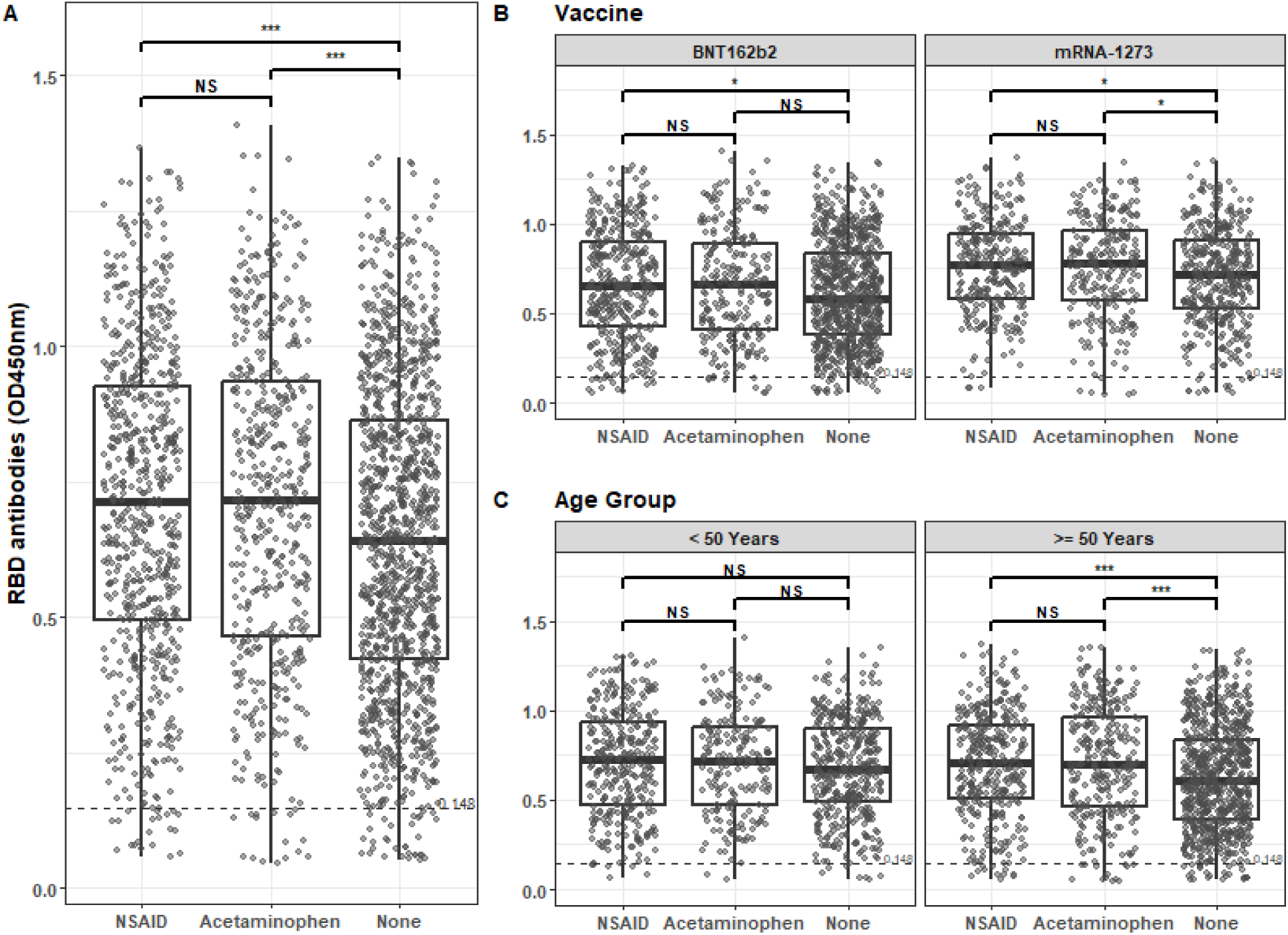
RBD-specific antibody levels post-vaccination. Pairwise means of RBD ELISA OD450 by pain reliever group (A), additionally stratified by vaccine (B) and age (C) were compared using one-way analysis variance. Pairwise differences were compared using two-sample t-test statistics with p-values adjustment using Tukey honestly significant difference (HSD). Significance levels: * p-value < 0.05; ** p-value < 0.01; *** p-value < 0.001.

**Table 1.** Demographics.

|  | NSAID<sup>1</sup><br>(N=679) | Acetaminophen<sup>2</sup><br>(N=491) | No Pain Medication<br>(N=1184) | Overall<br>(N=2354) |
| --- | --- | --- | --- | --- |
| **Age (years)** |  |  |  |  |
| Mean (SD) | 50.4 (15.9) | 53.4 (15.4) | 55.1 (16.6) | 53.4 (16.3) |
| Median [Min, Max] | 51.2 [18.2, 99.0] | 54.6 [18.0, 91.5] | 57.9 [18.0, 92.6] | 54.9 [18.0, 99.0] |
| **Sex at Birth** |  |  |  |  |
| Female | 488 (71.9%) | 367 (74.7%) | 693 (58.5%) | 1548 (65.8%) |
| Male | 185 (27.2%) | 123 (25.1%) | 491 (41.5%) | 799 (33.9%) |
| Missing | 6 (0.9%) | 1 (0.2%) | 0 (0%) | 7 (0.3%) |
| **Race/Ethnicity** |  |  |  |  |
| Hispanic or Latino | 108 (15.9%) | 98 (20.0%) | 173 (14.6%) | 379 (16.1%) |
| Non-Hispanic Black | 11 (1.6%) | 6 (1.2%) | 11 (0.9%) | 28 (1.2%) |
| Non-Hispanic Other | 64 (9.4%) | 46 (9.4%) | 119 (10.1%) | 229 (9.7%) |
| Non-Hispanic White | 496 (73.0%) | 341 (69.5%) | 881 (74.4%) | 1718 (73.0%) |
| **RBD (OD)** |  |  |  |  |
| Mean (SD) | 0.702 (0.292) | 0.708 (0.299) | 0.643 (0.285) | 0.673 (0.291) |
| Median [Min, Max] | 0.714 [0.0570, 1.37] | 0.717 [0.0470, 1.41] | 0.641 [0.0520, 1.35] | 0.676 [0.0470, 1.41] |
| **Serostatus** |  |  |  |  |
| Positive | 644 (94.8%) | 463 (94.3%) | 1121 (94.7%) | 2228 (94.6%) |
| Negative | 35 (5.2%) | 28 (5.7%) | 63 (5.3%) | 126 (5.4%) |

<sup>1</sup>e.g. Ibuprofen, Naproxen, Aspirin<sup>2</sup>e.g. Tylenol

Twenty-eight percent of the participants that completed the survey took NSAID pain relievers, compared to 21% taking acetaminophen and 50% not taking any pain medication. The most common vaccine-induced side effects were fatigue, muscle aches and headaches, and were higher in those taking NSAIDs and acetaminophen compared to those not taking any pain relievers (**Fig. 2A**). Logistic regression analyses showed that there was a statistically significantly higher proportion of participants taking pain medication that experienced fatigue and muscle aches, particularly between those taking NSAIDs and those not taking pain medication (p < 0.0001 for both vaccine side effects). We also observed a statistically higher proportion of NSAID users reporting headaches, compared to those not taking pain medication and lower than those taking acetaminophen (p < 0.0088). There were other side effects that were not captured on the survey (denoted as “other”), which were most common among those not taking any pain medication (**Fig. 2A**, 23.8%).

**Figure 2.**
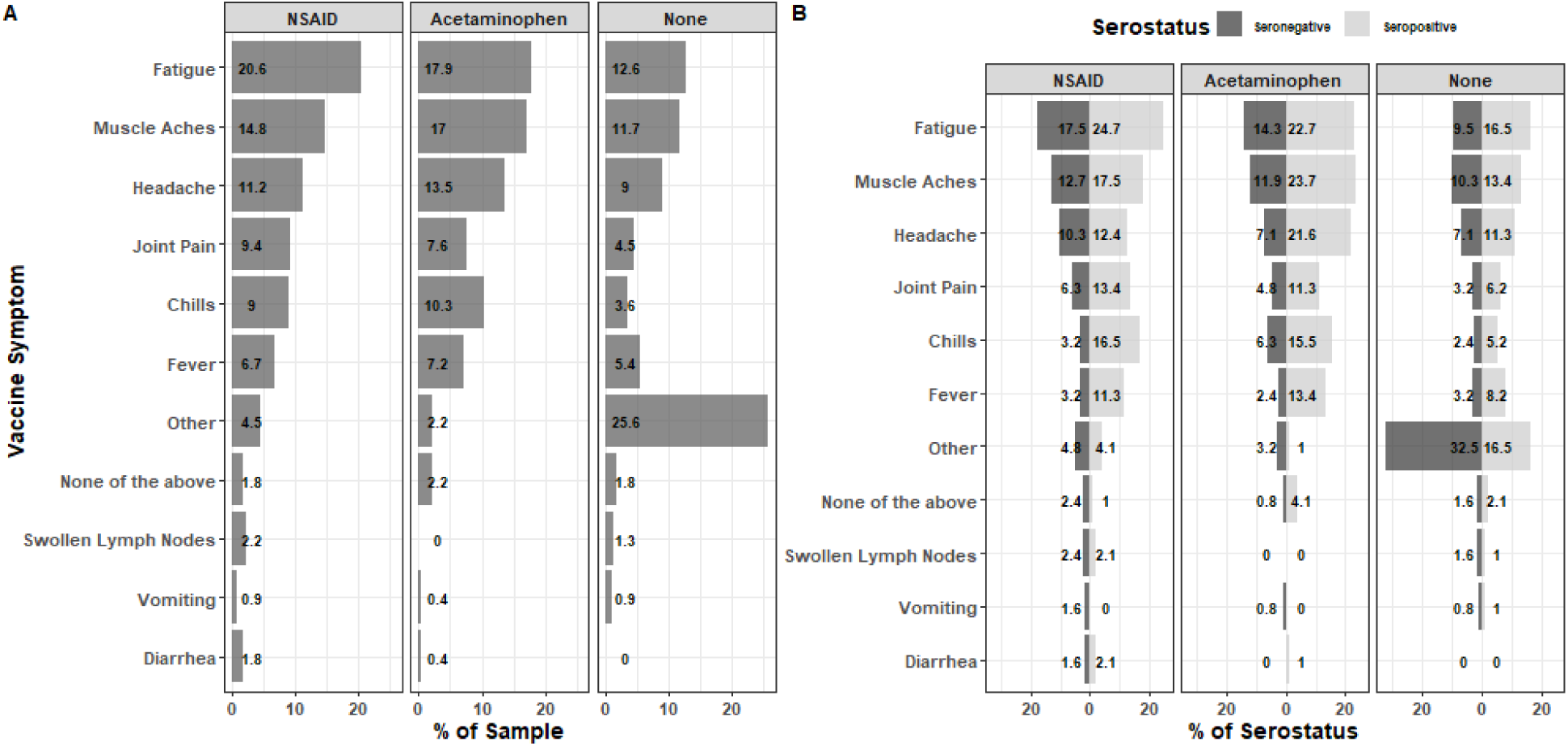
Rates of symptoms post-vaccination. Percent of participants experiencing each symptom (A) as well as stratified by serostatus (B). Logistic regression analysis was used to examine the difference between proportion of symptoms and pain reliever use. There was a statistically significant difference in the proportion of participants experiencing fatigue and muscle aches between NSAID users and those not taking pain medication (p < 0.0001 for both). We also observed a statistically higher proportion of headaches reported by NSAID users, compared to those not taking pain medication and lower than those taking acetaminophen (p < 0.0088).

While 95% of participants were scored as diagnostically seropositive after vaccination, a small fraction of individuals (126/2,354) fell below the seropositive threshold, based on RBD and S2 antibody levels (see Methods section for details). The percentage of participants reporting fatigue, muscle aches, headaches, joint pain, chills, and/or fever were greater in those who tested seropositive relative to those who were diagnostically seronegative (**Fig. 2B**). To further examine these relationships, we compared antibody levels in the subsets of participants who reported fatigue, muscle aches, and/or headaches across analgesic use categories (**Fig. 3A-C**). We observed no statistically significant differences in RBD-specific antibody levels as a function of analgesic use when participants were matched for these symptoms (**Fig. 3A-C**). These data suggest that inflammation and adverse events, rather than analgesic use per se, are associated with elevated antibodies.

**Figure 3.**
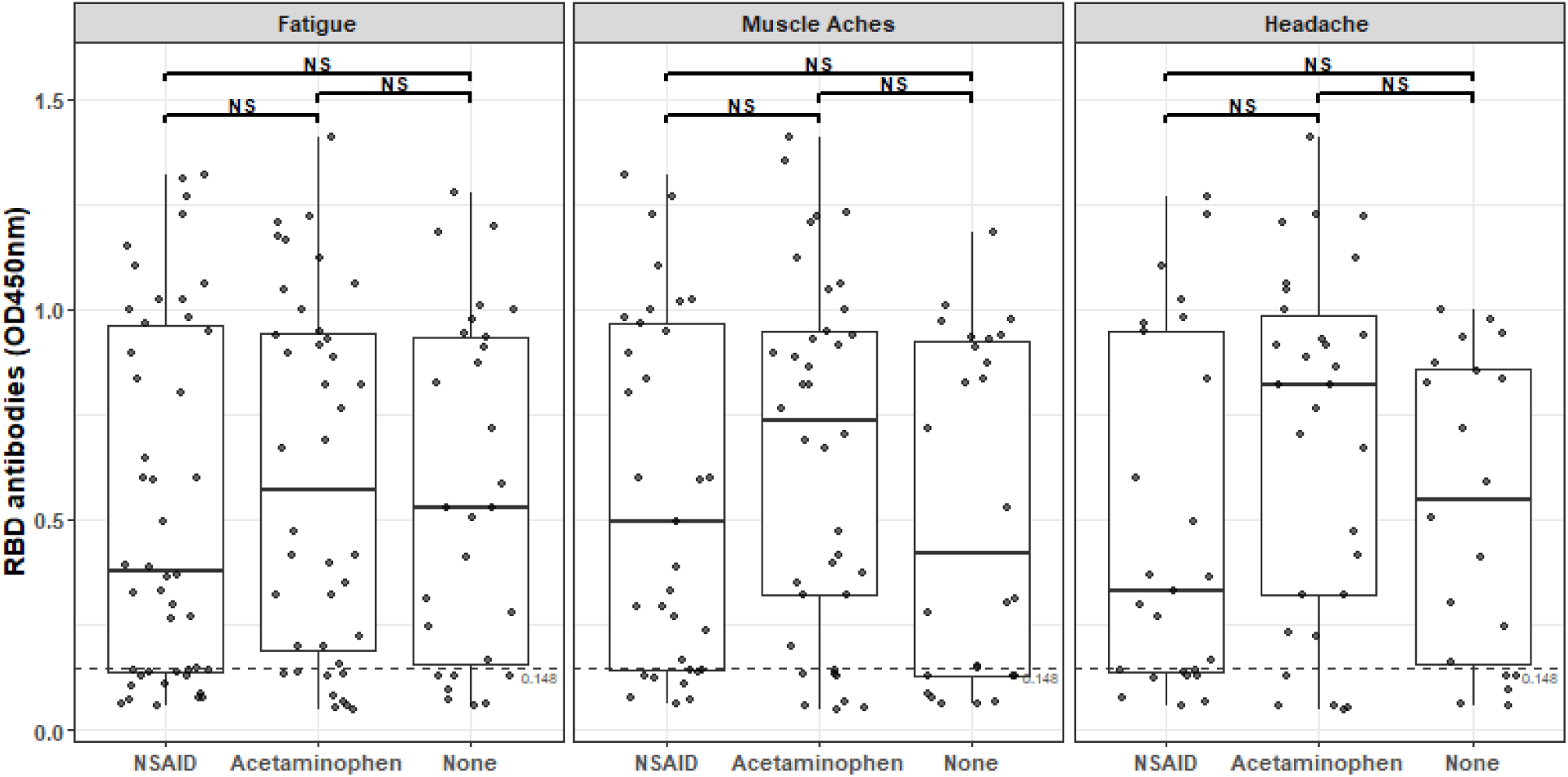
RBD-specific antibody levels as a function of symptoms and analgesic use. Pairwise means of RBD ELISA OD450 by pain reliever group for the top three vaccine induced symptoms. Pairwise differences were compared using two-sample t-test statistics with p-values adjustment using Tukey honestly significant difference (HSD).

## Discussion

Despite clear effectiveness against symptomatic infections and severe disease (10–12, 22), the uptake of both the original mRNA COVID-19 vaccines and updated boosters in the US has been lower than desired (14). There are many reasons for this low uptake, but one clear factor is the reactogenicity of the mRNA vaccines (14). For adenovirus-based COVID-19 vaccines, antipyretic use does alleviate symptoms (23), and may do so as well for mRNA vaccines. Widespread recommendation of their use after mRNA-vaccination may thus improve booster uptake.

Unlike animal models of SARS-CoV-2 infections (17), several prior studies observed no detriment to post-vaccination antibody responses by analgesic use, but differed on whether their use was in fact associated with elevated titers (18–20). A causal positive relationship between analgesic use and antibody levels would be unexpected, but in theory might be explained by the suppression of inflammation that is detrimental to B cell responses (24–30). Alternatively, several other studies reported positive associations between post-vaccination symptoms and antibody levels (31–34), which, biologically, seem more likely to be causally related. Since analgesic use is more common in those who experience symptoms, their association with antibody titers might simply be a correlative marker of these adverse events. By examining each of these parameters in a relatively large cohort, our data confirmed that analgesic use and certain symptoms correlate with elevated antibody levels. This association held across mRNA vaccines, age groups, and class of analgesics. However, a causal relationship between painkillers and antibody levels seems unlikely, as analgesic use did not obviously impact antibody responses independently of symptoms, positively or negatively.

Prior studies on SARS-CoV-2 infections showed clear positive associations of COVID-19 disease severity and inflammation with antibody levels (35), though the quality of the antibodies might be negatively impacted with excessive inflammation (30). It may be that for vaccines, inflammation and associated side effects are also causally linked to antibody titers. As one possible mechanistic explanation, certain HLA-haplotypes are associated with adverse events after vaccination (36). These haplotypes may also be associated with improved CD4+ T cell help for B cell responses and resultant antibody levels.

Though our data support widespread recommendations of analgesic use to mitigate post-COVID-19 vaccination side effects, current public health campaigns may encourage contemporaneous receipt of influenza and respiratory syncytial virus (RSV) vaccines. While the impact of analgesics on influenza vaccine responses has been reported to be minimal (37), their impact on RSV vaccine responses remains unknown.

## Limitations

Our study did not examine other parameters of immunity, such as memory B and T cells. Moreover, though our semi-quantitative RBD antibody assays correlate reasonably well with neutralizing antibodies (21), we did not directly measure antibody quality or function here.

## Data Availability

All data produced in the present study are available upon reasonable request to the authors

## Acknowledgements

We thank the volunteers across the State of Arizona who participated in this study. We thank the many members of the University of Arizona Antibody Testing Initiative for regulatory, legal, logistical, information technology, and communications support. We thank members of the University of Arizona Genetics Core for performing serological assays.

## References

1. Polack, F. P., S. J. Thomas, N. Kitchin, J. Absalon, A. Gurtman, S. Lockhart, J. L. Perez, G. Pérez Marc, E. D. Moreira, C. Zerbini, R. Bailey, K. A. Swanson, S. Roychoudhury, K. Koury, P. Li, W. V. Kalina, D. Cooper, R. W. Frenck, L. L. Hammitt, Ö. Türeci, H. Nell, A. Schaefer, S. Ünal, D. B. Tresnan, S. Mather, P. R. Dormitzer, U. Şahin, K. U. Jansen, W. C. Gruber, and C4591001 Clinical Trial Group. 2020. Safety and Efficacy of the BNT162b2 mRNA Covid-19 Vaccine. N Engl J Med 383: 2603–2615.

2. Baden, L. R., H. M. El Sahly, B. Essink, K. Kotloff, S. Frey, R. Novak, D. Diemert, S. A. Spector, N. Rouphael, C. B. Creech, J. McGettigan, S. Khetan, N. Segall, J. Solis, A. Brosz, C. Fierro, H. Schwartz, K. Neuzil, L. Corey, P. Gilbert, H. Janes, D. Follmann, M. Marovich, J. Mascola, L. Polakowski, J. Ledgerwood, B. S. Graham, H. Bennett, R. Pajon, C. Knightly, B. Leav, W. Deng, H. Zhou, S. Han, M. Ivarsson, J. Miller, T. Zaks, and COVE Study Group. 2021. Efficacy and Safety of the mRNA-1273 SARS-CoV-2 Vaccine. N Engl J Med 384: 403–416.

3. Collie, S., J. Champion, H. Moultrie, L.-G. Bekker, and G. Gray. 2021. Effectiveness of BNT162b2 Vaccine against Omicron Variant in South Africa. N Engl J Med.

4. Chalkias, S., J. Whatley, F. Eder, B. Essink, S. Khetan, P. Bradley, A. Brosz, N. McGhee, J. E. Tomassini, X. Chen, X. Zhao, A. Sutherland, X. Shen, B. Girard, D. K. Edwards, J. Feng, H. Zhou, S. Walsh, D. C. Montefiori, L. R. Baden, J. M. Miller, and R. Das. 2022. Safety and Immunogenicity of Omicron BA.4/BA.5 Bivalent Vaccine Against Covid-19. 2022.12.11.22283166.

5. Zou, J., C. Kurhade, S. Patel, N. Kitchin, K. Tompkins, M. Cutler, D. Cooper, Q. Yang, H. Cai, A. Muik, Y. Zhang, D.-Y. Lee, U. Sahin, A. S. Anderson, W. C. Gruber, X. Xie, K. A. Swanson, and P.-Y. Shi. 2022. Improved Neutralization of Omicron BA.4/5, BA.4.6, BA.2.75.2, BQ.1.1, and XBB.1 with Bivalent BA.4/5 Vaccine. 2022.11.17.516898.

6. Davis-Gardner, M. E., L. Lai, B. Wali, H. Samaha, D. Solis, M. Lee, A. Porter-Morrison, I. T. Hentenaar, F. Yamamoto, S. Godbole, Y. Liu, D. C. Douek, F. E.-H. Lee, N. Rouphael, A. Moreno, B. A. Pinsky, and M. S. Suthar. 2023. Neutralization against BA.2.75.2, BQ.1.1, and XBB from mRNA Bivalent Booster. New England Journal of Medicine 388: 183–185.

7. Kurhade, C., J. Zou, H. Xia, M. Liu, H. C. Chang, P. Ren, X. Xie, and P.-Y. Shi. 2022. Low neutralization of SARS-CoV-2 Omicron BA.2.75.2, BQ.1.1 and XBB.1 by parental mRNA vaccine or a BA.5 bivalent booster. Nat Med 1–4.

8. Zou, J., C. Kurhade, S. Patel, N. Kitchin, K. Tompkins, M. Cutler, D. Cooper, Q. Yang, H. Cai, A. Muik, Y. Zhang, D.-Y. Lee, U. Şahin, A. S. Anderson, W. C. Gruber, X. Xie, K. A. Swanson, and P.-Y. Shi. 2023. Neutralization of BA.4–BA.5, BA.4.6, BA.2.75.2, BQ.1.1, and XBB.1 with Bivalent Vaccine. New England Journal of Medicine 0: null.

9. Lin, D.-Y., Y. Xu, Y. Gu, D. Zeng, B. Wheeler, H. Young, S. K. Sunny, and Z. Moore. 2023. Effectiveness of Bivalent Boosters against Severe Omicron Infection. New England Journal of Medicine 0: null.

10. Link-Gelles, R. 2022. Effectiveness of Bivalent mRNA Vaccines in Preventing Symptomatic SARS-CoV-2 Infection — Increasing Community Access to Testing Program, United States, September–November 2022. MMWR Morb Mortal Wkly Rep 71.

11. Tenforde, M. W. 2022. Early Estimates of Bivalent mRNA Vaccine Effectiveness in Preventing COVID-19–Associated Emergency Department or Urgent Care Encounters and Hospitalizations Among Immunocompetent Adults — VISION Network, Nine States, September–November 2022. MMWR Morb Mortal Wkly Rep 71.

12. Surie, D. 2022. Early Estimates of Bivalent mRNA Vaccine Effectiveness in Preventing COVID-19–Associated Hospitalization Among Immunocompetent Adults Aged ≥65 Years — IVY Network, 18 States, September 8–November 30, 2022. MMWR Morb Mortal Wkly Rep 71.

13. 2022. COVID-19 Vaccination Coverage and Vaccine Confidence Among Adults | CDC.

14. Sinclair, A. H. 2023. Reasons for Receiving or Not Receiving Bivalent COVID-19 Booster Vaccinations Among Adults — United States, November 1–December 10, 2022. MMWR Morb Mortal Wkly Rep 72.

15. Walsh, E. E., R. W. Frenck, A. R. Falsey, N. Kitchin, J. Absalon, A. Gurtman, S. Lockhart, K. Neuzil, M. J. Mulligan, R. Bailey, K. A. Swanson, P. Li, K. Koury, W. Kalina, D. Cooper, C. Fontes-Garfias, P.-Y. Shi, Ö. Türeci, K. R. Tompkins, K. E. Lyke, V. Raabe, P. R. Dormitzer, K. U. Jansen, U. Şahin, and W. C. Gruber. 2020. Safety and Immunogenicity of Two RNA-Based Covid-19 Vaccine Candidates. New England Journal of Medicine 383: 2439–2450.

16. Jackson, L. A., E. J. Anderson, N. G. Rouphael, P. C. Roberts, M. Makhene, R. N. Coler, M. P. McCullough, J. D. Chappell, M. R. Denison, L. J. Stevens, A. J. Pruijssers, A. McDermott, B. Flach, N. A. Doria-Rose, K. S. Corbett, K. M. Morabito, S. O’Dell, S. D. Schmidt, P. A. Swanson, M. Padilla, J. R. Mascola, K. M. Neuzil, H. Bennett, W. Sun, E. Peters, M. Makowski, J. Albert, K. Cross, W. Buchanan, R. Pikaart-Tautges, J. E. Ledgerwood, B. S. Graham, and J. H. Beigel. 2020. An mRNA Vaccine against SARS-CoV-2 — Preliminary Report. New England Journal of Medicine 383: 1920–1931.

17. Chen, J. S., M. M. Alfajaro, R. D. Chow, J. Wei, R. B. Filler, S. C. Eisenbarth, and C. B. Wilen. 2021. Non-steroidal anti-inflammatory drugs dampen the cytokine and antibody response to SARS-CoV-2 infection. J Virol JVI.00014–21.

18. Tani, N., H. Ikematsu, T. Goto, K. Gondo, T. Inoue, Y. Yanagihara, Y. Kurata, R. Oishi, J. Minami, K. Onozawa, S. Nagano, H. Kuwano, K. Akashi, N. Shimono, and Y. Chong. 2022. Correlation of Postvaccination Fever With Specific Antibody Response to Severe Acute Respiratory Syndrome Coronavirus 2 BNT162b2 Booster and No Significant Influence of Antipyretic Medication. Open Forum Infectious Diseases 9: ofac493.

19. Tani, N., Y. Chong, Y. Kurata, K. Gondo, R. Oishi, T. Goto, J. Minami, K. Onozawa, S. Nagano, N. Shimono, H. Ikematsu, and H. Kuwano. 2022. Relation of fever intensity and antipyretic use with specific antibody response after two doses of the BNT162b2 mRNA vaccine. Vaccine 40: 2062–2067.

20. Choi, M. J., J. Y. Heo, Y. B. Seo, Y. K. Yoon, J. W. Sohn, J. Y. Noh, H. J. Cheong, W. J. Kim, J. Choi, Y. J. Lee, H. W. Lee, S. S. Kim, B. Kim, and J. Y. Song. 2023. Predictive Value of Reactogenicity for Anti-SARS-CoV-2 Antibody Response in mRNA-1273 Recipients: A Multicenter Prospective Cohort Study. Vaccines 11: 120.

21. Ripperger, T. J., J. L. Uhrlaub, M. Watanabe, R. Wong, Y. Castaneda, H. A. Pizzato, M. R. Thompson, C. Bradshaw, C. C. Weinkauf, C. Bime, H. L. Erickson, K. Knox, B. Bixby, S. Parthasarathy, S. Chaudhary, B. Natt, E. Cristan, T. El Aini, F. Rischard, J. Campion, M. Chopra, M. Insel, A. Sam, J. L. Knepler, A. P. Capaldi, C. M. Spier, M. D. Dake, T. Edwards, M. E. Kaplan, S. J. Scott, C. Hypes, J. Mosier, D. T. Harris, B. J. LaFleur, R. Sprissler, J. Nikolich-Žugich, and D. Bhattacharya. 2020. Orthogonal SARS-CoV-2 Serological Assays Enable Surveillance of Low-Prevalence Communities and Reveal Durable Humoral Immunity. Immunity 53: 925–933.e4.

22. McMenamin, M. E., J. Nealon, Y. Lin, J. Y. Wong, J. K. Cheung, E. H. Y. Lau, P. Wu, G. M. Leung, and B. J. Cowling. 2022. Vaccine effectiveness of one, two, and three doses of BNT162b2 and CoronaVac against COVID-19 in Hong Kong: a population-based observational study. Lancet Infect Dis 22: 1435–1443.

23. Folegatti, P. M., K. J. Ewer, P. K. Aley, B. Angus, S. Becker, S. Belij-Rammerstorfer, D. Bellamy, S. Bibi, M. Bittaye, E. A. Clutterbuck, C. Dold, S. N. Faust, A. Finn, A. L. Flaxman, B. Hallis, P. Heath, D. Jenkin, R. Lazarus, R. Makinson, A. M. Minassian, K. M. Pollock, M. Ramasamy, H. Robinson, M. Snape, R. Tarrant, M. Voysey, C. Green, A. D. Douglas, A. V. S. Hill, T. Lambe, S. C. Gilbert, A. J. Pollard, J. Aboagye, K. Adams, A. Ali, E. Allen, J. L. Allison, R. Anslow, E. H. Arbe-Barnes, G. Babbage, K. Baillie, M. Baker, N. Baker, P. Baker, I. Baleanu, J. Ballaminut, E. Barnes, J. Barrett, L. Bates, A. Batten, K. Beadon, R. Beckley, E. Berrie, L. Berry, A. Beveridge, K. R. Bewley, E. M. Bijker, T. Bingham, L. Blackwell, C. L. Blundell, E. Bolam, E. Boland, N. Borthwick, T. Bower, A. Boyd, T. Brenner, P. D. Bright, C. Brown-O’Sullivan, E. Brunt, J. Burbage, S. Burge, K. R. Buttigieg, N. Byard, I. C. Puig, A. Calvert, S. Camara, M. Cao, F. Cappuccini, M. Carr, M. W. Carroll, V. Carter, K. Cathie, R. J. Challis, S. Charlton, I. Chelysheva, J.-S. Cho, P. Cicconi, L. Cifuentes, H. Clark, E. Clark, T. Cole, R. Colin-Jones, C. P. Conlon, A. Cook, N. S. Coombes, R. Cooper, C. A. Cosgrove, K. Coy, W. E. M. Crocker, C. J. Cunningham, A. E. Damratoski, L. Dando, M. S. Datoo, H. Davies, H. D. Graaf, T. Demissie, C. D. Maso, I. Dietrich, T. Dong, F. R. Donnellan, N. Douglas, C. Downing, J. Drake, R. Drake-Brockman, R. E. Drury, S. J. Dunachie, N. J. Edwards, F. D. L. Edwards, C. J. Edwards, S. C. Elias, M. J. Elmore, K. R. W. Emary, M. R. English, S. Fagerbrink, S. Felle, S. Feng, S. Field, C. Fixmer, C. Fletcher, K. J. Ford, J. Fowler, P. Fox, E. Francis, J. Frater, J. Furze, M. Fuskova, E. Galiza, D. Gbesemete, C. Gilbride, K. Godwin, G. Gorini, L. Goulston, C. Grabau, L. Gracie, Z. Gray, L. B. Guthrie, M. Hackett, S. Halwe, E. Hamilton, J. Hamlyn, B. Hanumunthadu, I. Harding, S. A. Harris, A. Harris, D. Harrison, C. Harrison, T. C. Hart, L. Haskell, S. Hawkins, I. Head, J. A. Henry, J. Hill, S. H. C. Hodgson, M. M. Hou, E. Howe, N. Howell, C. Hutlin, S. Ikram, C. Isitt, P. Iveson, S. Jackson, F. Jackson, S. W. James, M. Jenkins, E. Jones, K. Jones, C. E. Jones, B. Jones, R. Kailath, K. Karampatsas, J. Keen, S. Kelly, D. Kelly, D. Kerr, S. Kerridge, L. Khan, U. Khan, A. Killen, J. Kinch, T. B. King, L. King, J. King, L. Kingham-Page, P. Klenerman, F. Knapper, J. C. Knight, D. Knott, S. Koleva, A. Kupke, C. W. Larkworthy, J. P. J. Larwood, A. Laskey, A. M. Lawrie, A. Lee, K. Y. N. Lee, E. A. Lees, H. Legge, A. Lelliott, N.-M. Lemm, A. M. Lias, A. Linder, S. Lipworth, X. Liu, S. Liu, R. L. Ramon, M. Lwin, F. Mabesa, M. Madhavan, G. Mallett, K. Mansatta, I. Marcal, S. Marinou, E. Marlow, J. L. Marshall, J. Martin, J. McEwan, L. McInroy, G. Meddaugh, A. J. Mentzer, N. Mirtorabi, M. Moore, E. Moran, E. Morey, V. Morgan, S. J. Morris, H. Morrison, G. Morshead, R. Morter, Y. F. Mujadidi, J. Muller, T. Munera-Huertas, A. Munro, A. Munro, S. Murphy, V. J. Munster, P. Mweu, A. Noé, F. L. Nugent, E. Nuthall, K. O’Brien, D. O’Connor, B. Oguti, J. L. Oliver, C. Oliveira, P. J. O’Reilly, M. Osborn, P. Osborne, C. Owen, D. Owens, N. Owino, M. Pacurar, K. Parker, H. Parracho, M. Patrick-Smith, V. Payne, J. Pearce, Y. Peng, M. P. P. Alvarez, J. Perring, K. Pfafferott, D. Pipini, E. Plested, H. Pluess-Hall, K. Pollock, I. Poulton, L. Presland, S. Provstgaard-Morys, D. Pulido, K. Radia, F. R. Lopez, J. Rand, H. Ratcliffe, T. Rawlinson, S. Rhead, A. Riddell, A. J. Ritchie, H. Roberts, J. Robson, S. Roche, C. Rohde, C. S. Rollier, R. Romani, I. Rudiansyah, S. Saich, S. Sajjad, S. Salvador, L. S. Riera, H. Sanders, K. Sanders, S. Sapaun, C. Sayce, E. Schofield, G. Screaton, B. Selby, C. Semple, H. R. Sharpe, I. Shaik, A. Shea, H. Shelton, S. Silk, L. Silva-Reyes, D. T. Skelly, H. Smee, C. C. Smith, D. J. Smith, R. Song, A. J. Spencer, E. Stafford, A. Steele, E. Stefanova, L. Stockdale, A. Szigeti, A. Tahiri-Alaoui, M. Tait, H. Talbot, R. Tanner, I. J. Taylor, V. Taylor, R. T. W. Naude, N. Thakur, Y. Themistocleous, A. Themistocleous, M. Thomas, T. M. Thomas, A. Thompson, S. Thomson-Hill, J. Tomlins, S. Tonks, J. Towner, N. Tran, J. A. Tree, A. Truby, K. Turkentine, C. Turner, N. Turner, S. Turner, T. Tuthill, M. Ulaszewska, R. Varughese, N. V. Doremalen, K. Veighey, M. K. Verheul, I. Vichos, E. Vitale, L. Walker, M. E. E. Watson, B. Welham, J. Wheat, C. White, R. White, A. T. Worth, D. Wright, S. Wright, X. L. Yao, and Y. Yau. 2020. Safety and immunogenicity of the ChAdOx1 nCoV-19 vaccine against SARS-CoV-2: a preliminary report of a phase 1/2, single-blind, randomised controlled trial. The Lancet 396: 467–478.

24. Biram, A., J. Liu, H. Hezroni, N. Davidzohn, D. Schmiedel, E. Khatib-Massalha, M. Haddad, A. Grenov, S. Lebon, T. M. Salame, N. Dezorella, D. Hoffman, P. Abou Karam, M. Biton, T. Lapidot, M. Bemark, R. Avraham, S. Jung, and Z. Shulman. 2022. Bacterial infection disrupts established germinal center reactions through monocyte recruitment and impaired metabolic adaptation. Immunity 55: 442–458.e8.

25. Popescu, M., B. Cabrera-Martinez, and G. M. Winslow. 2019. TNF-α Contributes to Lymphoid Tissue Disorganization and Germinal Center B Cell Suppression during Intracellular Bacterial Infection. J Immunol 203: 2415–2424.

26. Keitany, G. J., K. S. Kim, A. T. Krishnamurty, B. D. Hondowicz, W. O. Hahn, N. Dambrauskas, D. N. Sather, A. M. Vaughan, S. H. I. Kappe, and M. Pepper. 2016. Blood Stage Malaria Disrupts Humoral Immunity to the Pre-erythrocytic Stage Circumsporozoite Protein. Cell Rep 17: 3193–3205.

27. Rothaeusler, K., and N. Baumgarth. 2010. B cell fate decisions following influenza virus infection. Eur J Immunol 40: 366–377.

28. Ryg-Cornejo, V., L. J. Ioannidis, A. Ly, C. Y. Chiu, J. Tellier, D. L. Hill, S. P. Preston, M. Pellegrini, D. Yu, S. L. Nutt, A. Kallies, and D. S. Hansen. 2016. Severe Malaria Infections Impair Germinal Center Responses by Inhibiting T Follicular Helper Cell Differentiation. Cell Reports 14: 68–81.

29. Röltgen, K., S. C. A. Nielsen, O. Silva, S. F. Younes, M. Zaslavsky, C. Costales, F. Yang, O. F. Wirz, D. Solis, R. A. Hoh, A. Wang, P. S. Arunachalam, D. Colburg, S. Zhao, E. Haraguchi, A. S. Lee, M. M. Shah, M. Manohar, I. Chang, F. Gao, V. Mallajosyula, C. Li, J. Liu, M. J. Shoura, S. B. Sindher, E. Parsons, N. J. Dashdorj, N. D. Dashdorj, R. Monroe, G. E. Serrano, T. G. Beach, R. S. Chinthrajah, G. W. Charville, J. L. Wilbur, J. N. Wohlstadter, M. M. Davis, B. Pulendran, M. L. Troxell, G. B. Sigal, Y. Natkunam, B. A. Pinsky, K. C. Nadeau, and S. D. Boyd. 2022. Immune imprinting, breadth of variant recognition, and germinal center response in human SARS-CoV-2 infection and vaccination. Cell S0092-8674(22)00076–9.

30. Kaneko, N., H.-H. Kuo, J. Boucau, J. R. Farmer, H. Allard-Chamard, V. S. Mahajan, A. Piechocka-Trocha, K. Lefteri, M. Osborn, J. Bals, Y. C. Bartsch, N. Bonheur, T. M. Caradonna, J. Chevalier, F. Chowdhury, T. J. Diefenbach, K. Einkauf, J. Fallon, J. Feldman, K. K. Finn, P. Garcia-Broncano, C. A. Hartana, B. M. Hauser, C. Jiang, P. Kaplonek, M. Karpell, E. C. Koscher, X. Lian, H. Liu, J. Liu, N. L. Ly, A. R. Michell, Y. Rassadkina, K. Seiger, L. Sessa, S. Shin, N. Singh, W. Sun, X. Sun, H. J. Ticheli, M. T. Waring, A. L. Zhu, G. Alter, J. Z. Li, D. Lingwood, A. G. Schmidt, M. Lichterfeld, B. D. Walker, X. G. Yu, R. F. Padera, S. Pillai, and Massachusetts Consortium on Pathogen Readiness Specimen Working Group. 2020. Loss of Bcl-6-Expressing T Follicular Helper Cells and Germinal Centers in COVID-19. Cell 183: 143–157.e13.

31. Hermann, E. A., B. Lee, P. P. Balte, V. Xanthakis, B. D. Kirkpatrick, M. Cushman, and E. Oelsner. 2022. Association of Symptoms After COVID-19 Vaccination With Anti–SARS-CoV-2 Antibody Response in the Framingham Heart Study. JAMA Network Open 5: e2237908.

32. Peikert, A., B. L. Claggett, K. Kim, J. A. Udell, J. Joseph, A. S. Desai, M. E. Farkouh, S. M. Hegde, A. F. Hernandez, D. L. Bhatt, J. M. Gaziano, H. K. Talbot, C. Yancy, I. Anand, L. Mao, L. S. Cooper, S. D. Solomon, and O. Vardeny. 2023. Association of post-vaccination adverse reactions after influenza vaccine with mortality and cardiopulmonary outcomes in patients with high-risk cardiovascular disease: the INVESTED trial. European Journal of Heart Failure 25: 299–310.

33. Goel, R. R., S. A. Apostolidis, M. M. Painter, D. Mathew, A. Pattekar, O. Kuthuru, S. Gouma, P. Hicks, W. Meng, A. M. Rosenfeld, S. Dysinger, K. A. Lundgreen, L. Kuri-Cervantes, S. Adamski, A. Hicks, S. Korte, D. A. Oldridge, A. E. Baxter, J. R. Giles, M. E. Weirick, C. M. McAllister, J. Dougherty, S. Long, K. D’Andrea, J. T. Hamilton, M. R. Betts, E. T. L. Prak, P. Bates, S. E. Hensley, A. R. Greenplate, and E. J. Wherry. 2021. Distinct antibody and memory B cell responses in SARS-CoV-2 naïve and recovered individuals following mRNA vaccination. Science Immunology 6.

34. Dutcher, E. G., E. S. Epel, A. E. Mason, F. M. Hecht, J. E. Robinson, S. S. Drury, and A. A. Prather. 2023. The more symptoms the better? Covid-19 vaccine side effects and long-term neutralizing antibody response. 2023.09.26.23296186.

35. Long, Q.-X., X.-J. Tang, Q.-L. Shi, Q. Li, H.-J. Deng, J. Yuan, J.-L. Hu, W. Xu, Y. Zhang, F.-J. Lv, K. Su, F. Zhang, J. Gong, B. Wu, X.-M. Liu, J.-J. Li, J.-F. Qiu, J. Chen, and A.-L. Huang. 2020. Clinical and immunological assessment of asymptomatic SARS-CoV-2 infections. Nat. Med. 10.1038/s41591-020-0965-6.

36. Bolze, A., I. Neveux, K. M. Schiabor Barrett, S. White, M. Isaksson, S. Dabe, W. Lee, J. J. Grzymski, N. L. Washington, and E. T. Cirulli. 2022. HLA-A∗03:01 is associated with increased risk of fever, chills, and stronger side effects from Pfizer-BioNTech COVID-19 vaccination. HGG Adv 3: 100084.

37. Goodwin, J. S., D. S. Selinger, R. P. Messner, and W. P. Reed. 1978. Effect of indomethacin in vivo on humoral and cellular immunity in humans. Infect Immun 19: 430–433.

